# Improved correction of anatomical distortion in diffusion-weighted prostate MRI

**DOI:** 10.1101/2024.03.26.24304935

**Authors:** Joshua S Kim, Christopher C Conlin, Mark A Anderson, Tristan Barrett, Son Do, Leonidas Georgiou, Mukesh G Harisinghani, Michael A Liss, Ryan Manger, Daniel JA Margolis, Robert M Marks, Paul M Murphy, Nabih Nakrour, Michael Ohliger, Todd Pawlicki, Rebecca Rakow-Penner, Mariluz Rojo Domingo, Jessica Scholey, Steven N Seyedin, Yuze Song, Natasha Wehrli, Sean Woolen, Constantinos Zamboglou, Tyler M Seibert, Anders M Dale

## Abstract

**Purpose:** *B0*-inhomogeneity distortion limits the utility of diffusion weighted imaging (DWI) for prostate cancer (PCa) lesion targeting for focal radiotherapy. Distortion correction methods like the Reversed Polarity Gradient (RPG) method and FSL-topup struggle due to signal-intensity changes from voxel stretch/overlap. We propose a technique called multi-b RPG (mRPG) for correcting *B_0_*-inhomogeneity distortions that eliminates confounding signal-intensity changes and demonstrate that it outperforms alternative methods.

**Methods and Materials:** mRPG employs DWI volumes of opposite phase-encoding polarity acquired at different *b*-values. Signal-intensity changes from voxel stretch/overlap are eliminated by normalizing the volumes by the sum across all *b*-values. Tissue deformation is then estimated by registering normalized volumes of opposite phase-encoding polarity.

Patients with known/suspected PCa were included if scanned with a protocol including two multi-*b*-value DWI series with opposite phase-encoding polarity. Degree of anatomic distortion was quantified by the Pearson correlation between opposite-polarity volumes; less distortion yields higher correlation. Correlation coefficients were computed for DWI and ADC volumes of each patient, before and after correction by RPG, topup, and mRPG. Absolute tissue displacement was measured along the posterior prostate border and within PCa lesions.

**Results:** 1225 patients were available for analysis. mRPG yielded significant improvements in median correlation (*p*<0.001) between opposite-polarity DWI volumes (0.90 vs 0.83 uncorrected) and ADC volumes (0.78 vs 0.71). The increase achieved with mRPG was significantly larger (*p*<0.001) than with either RPG or topup on DWI (0.90 vs 0.84 with RPG; 0.81 with topup) or ADC (0.78 vs 0.71 with RPG; 0.67 with topup).

Tissue displacement during correction ranged from 0.5–13.8mm along the posterior border of the prostate and 0.5–7.5mm within PCa lesions. Displacement ≥4mm was observed in 30% of patients.

**Conclusions:** mRPG is a feasible method for correcting *B_0_*-inhomogeneity distortions in prostate DWI that avoids drawbacks of alternative methods to achieve better distortion correction performance.

## Introduction

Diffusion-weighted imaging (DWI) is important for identifying tumors in patients with suspected prostate cancer. In addition to its longstanding diagnostic utility as part of the Prostate Imaging Reporting and Data System (PI-RADS) (1), DWI and the apparent diffusion coefficient (ADC) derived from DWI have proven to be valuable aids for accurately targeting prostate tumors with radiation therapy (2). However, DWI data acquired with standard echo-planar imaging (EPI) trajectories are prone to distortion artifacts that arise from inhomogeneities in the static magnetic field (*B_0_*). The tissue-air interface where the prostate meets the rectum is a common source of *B_0_* inhomogeneities that cause considerable distortion of the local anatomy, which can obscure the precise location of a lesion and limits accuracy of lesion targeting for focal radiotherapy (RT) boost (3).

*B_0_*-inhomogeneity distortion artifacts present as compression or expansion of tissue in the phase-encoding direction, with compressed regions displaying increased signal intensity from the additional spins (water protons) incorrectly localized to each voxel. Conversely, expanded regions show reduced signal intensity from the apparent decrease in spins within each voxel. These effects can be described mathematically according to Equation 1:

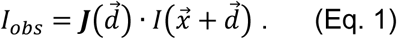

The signal intensity of the acquired image, *I*, is a function of the true voxel position 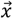, plus the tissue displacement 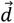 along the phase-encoding direction that results from distortion. Signal intensity is also scaled by the Jacobian ***J***, which is the partial derivative of tissue displacement along the phase-encoding direction. The Jacobian quantifies the amount of local compression or expansion of tissue and the resulting concentration or dilution of signal intensity in the final observed image, *I_obs_*.

Most clinical platforms do not routinely include any correction for these *B_0_*-inhomogeneity distortion effects, though methods have been proposed for correction. One common approach in the research setting involves the acquisition of two EPI images, typically without diffusion weighting (i.e., *b*=0 s/mm^2^) and with opposite phase-encoding polarity (hereafter arbitrarily referred to as the “forward” and “reverse” images). These images will show distortion artifacts that are equal in magnitude but in opposite directions, with tissue compression in one image and expansion in the other. The true position of a voxel of undistorted tissue will therefore be in the middle of its corresponding position on the two opposite-polarity images. Distortion correction techniques like the Reversed Polarity Gradient (RPG) method (3, 4) and FSL-topup (5, 6) use numerical optimization to estimate the tissue displacement field 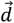 that minimizes the difference between forward and reverse images:

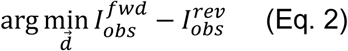

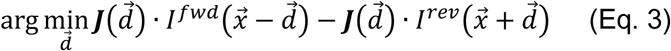

(This is a simplified description of the minimization problem, shown to introduce the theoretical underpinning of the approach. In practice, the objective function is defined to be quadratic and is subject to various constraints to improve optimization performance.) Anatomical distortion is then corrected by shifting the voxels of the original images by the estimated displacement field 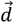.

Finding an accurate solution to Eq. 3 can be difficult, however, since the difference between forward and reverse images is influenced by the signal-intensity scaling factor, 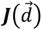—commonly referred to as the Jacobian Intensity Correction (JIC)—in addition to the spatial displacement of tissue. In this study, we propose an alternative, two-step approach for correcting *B_0_*-inhomogeneity distortions that first eliminates the signal-intensity scaling factor prior to estimating spatial displacement. We hypothesize that this two-step approach will improve the correction of anatomical distortion in prostate DWI compared to established techniques. Corrected DWI would overlay more accurately on anatomic *T_2_*-weighted images, thus potentially improving targeting of prostate lesions for focal RT boost or MRI-targeted biopsy.

## Methods and Materials

### Proposed distortion correction method (multi-*b* RPG)

The key observation underpinning the proposed method is that the Jacobian signal-intensity scaling factor 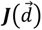 is only a function of tissue displacement; it does not depend on the *b*-value of the diffusion-weighted image. If images are acquired at two or more unique *b*-values, the Jacobian factor can be eliminated by normalizing an image by the sum over the different *b*-values:

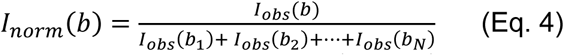

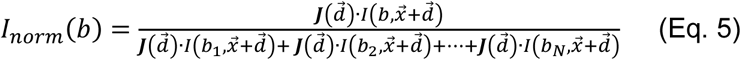

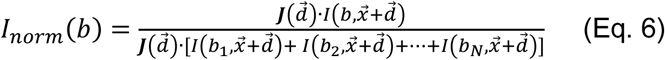

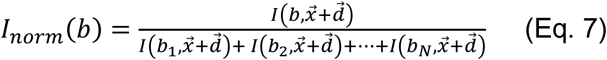

where *I_norm_*(*b*) is the normalized image computed from a DWI image acquired at any particular *b*-value *b*, and *I_obs_*(*b*_1_), *I_obs_*(*b*_2_), *I_obs_*(*b_N_*) are the images acquired at the 1^st^, 2^nd^, and N^th^ *b*-values, respectively.

If multiple unique *b*-values are acquired in both phase-encoding directions, the forward and reverse images can both be normalized. Substituting normalized images for the originals in Eqs. 2–3 simplifies the minimization problem by eliminating the Jacobian factor, i.e., the intensity changes that result from tissue compression or expansion. Estimating 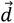 is therefore reduced to a much simpler image registration problem, for which any suitable algorithm may be used. In this study, registration was performed by shifting the forward and reverse volumes along the anterior-posterior axis and computing the amount of shift for each voxel that minimizes the difference between forward and reverse images (a detailed description of the registration algorithm is provided in the Supplementary Materials). This approach amounts to a multi-*b*-value adaptation of RPG, so we call it multi-*b* RPG (mRPG).

Figure 1 shows an example application of the proposed mRPG method to correct EPI distortion of prostate diffusion data. Prior to correction, the posterior prostate (abutting the rectum) is stretched in the forward image and compressed in the reverse, relative to the undistorted anatomy in the reference spin echo image. Signal enhancement of compressed tissue is readily apparent along the posterior edge of the prostate in the reverse *b*=0 s/mm^2^ image. Normalizing the original images by the sum across *b*-values eliminates the signal intensity changes resulting from tissue compression or expansion, yielding a much more uniform grey-level across the prostate (Figure 1B). Tissue displacement maps are then estimated by registering the normalized images, which are then applied to remove the original distortions. Following correction by mRPG, the posterior edge of the prostate in the diffusion images is better aligned with that of the undistorted reference image.

**Figure 1:**
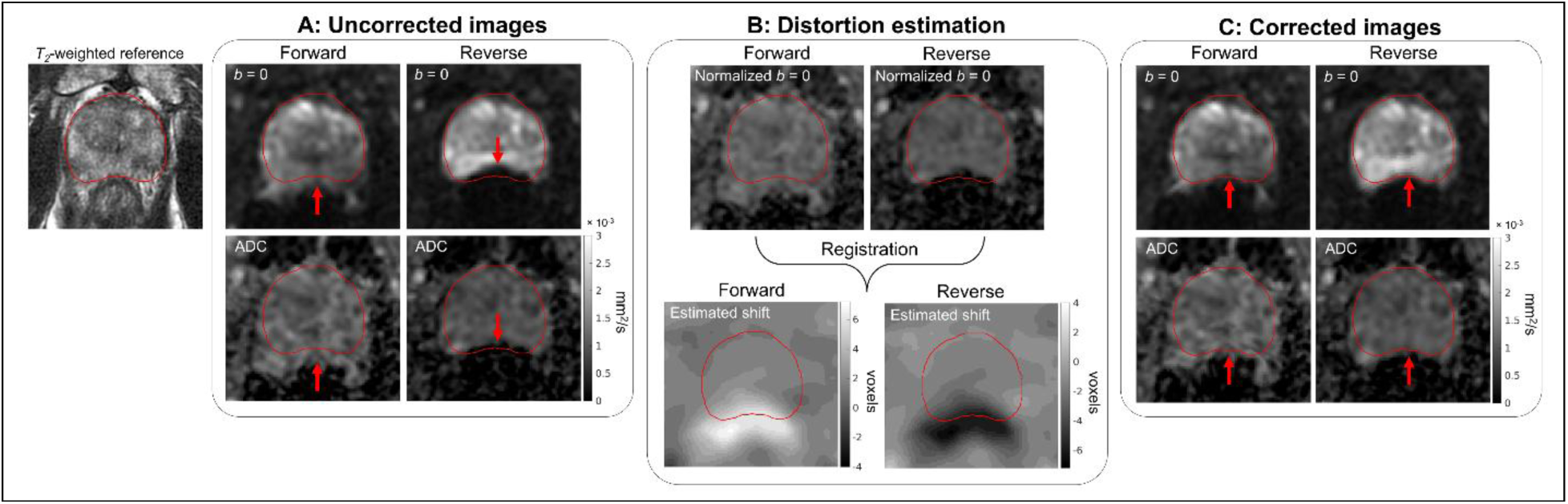
Example of mRPG distortion correction. An undistorted spin echo (*T2*-weighted) image is shown on the left, with the prostate outlined in red. A) Uncorrected diffusion data, with stretching of the posterior prostate in the forward direction and compression in the reverse (arrows). B) Registration of the normalized forward-and reverse-images yields the estimated voxel shifts. C) Corrected images after application of the shift maps in B, with reduced spatial distortion (arrows)

### Study participant enrollment

Four institutions participated in this multisite, multinational retrospective study: University of California San Diego (UCSD; primary site), University of California San Francisco (UCSF), University of Michigan (UM), and the German Oncology Center, Cyprus (GOC). Research activity was approved by the local Institutional Review Board (IRB) or Ethics Committee of each institution. At UCSD, UCSF, and Michigan, a waiver of consent was granted to access MRI exam data. At GOC, patients were enrolled after providing their written informed consent, as part of a separate, prospective trial that is approved by the local ethics committee (NCT07568613).

Patients were considered for enrollment if they underwent MRI for suspected or biopsy-proven prostate cancer between March 2023 and January 2026 at one of four participating institutions with a protocol that included a multi-*b*-value DWI acquisition as described below. Patients were excluded if they had metal hip implants or other devices that caused severe EPI-distortion artifacts.

### MRI acquisition and pre-processing

All MR imaging was performed on 3T clinical scanners using a 32-channel phased-array body coil placed over the pelvis. Four MRI scanner platforms were employed: SIGNA Premier from GE Healthcare and MAGNETOM Prisma, Vida, and Skyra from Siemens Healthineers. Acquisition parameters are summarized in Table 1. In addition to conventional images, two axial multi-*b*-value DWI volumes were acquired per patient, using EPI readouts with opposite phase-encoding polarity along the anterior-posterior axis; one acquired in the posterior-to-anterior direction and the other in the anterior-to-posterior direction (“forward” and “reverse” volumes, respectively). The forward and reverse volumes were otherwise identical and were typically acquired one immediately after the other. The full field of view was acquired using parallel imaging (ASSET on GE scanners, GRAPPA on Siemens) with an acceleration factor of 2. These additional images are included in routine clinical care at the four participating institutions. Water-selective excitation pulses were used for fat suppression. In a small subset of the patients, these DWI acquisitions were repeated an additional time within the same scanning session as part of a separate, prospective study with written consent (NCT04992728) (7). High resolution *T_2_*-weighted volumes without fat suppression were acquired for anatomical reference. This protocol was developed to support advanced quantitative DWI (2, 8–10) and is currently in clinical use at all four institutions participating in this study, as well as part of a number of prospective trials (NCT06579417, NCT05882253, NCT06990542) (11, 12).

**Table 1:** MRI acquisition details for the patients included in this study. Two axial DWI volumes were acquired per patient using the same diffusion-weighted spin-echo pulse sequence (default tensor), but with EPI readout trajectories of opposite phase-encoding polarity along the anterior-posterior axis. One was acquired in the posterior-to-anterior direction (“forward”) and the other in the anterior-to-posterior direction (“reverse”). Water-selective excitation pulses were used for fat signal suppression. The high-resolution *T2*-weighted (T2W) volume was acquired for anatomical reference using a fast spin-echo (FSE) pulse sequence without fat suppression.

|  | Acquisition time<br>(min:sec) | b-values (s/mm<sup>2</sup>)<br>[number of samples] | FOV (mm) | Matrix | Slices | Slice thickness<br>(mm) | TR (ms) | TE (ms) |
| --- | --- | --- | --- | --- | --- | --- | --- | --- |
| DWI forward | 3:25 | 0 [4], 100 [6], 800 [12],<br>1400 [12], 2500 [18] | 200×200 | 80×80 | 32 | 3 | 4500 | 80 |
| DWI reverse | 3:25 | 0 [4], 100 [6], 800 [12],<br>1400 [12], 2500 [18] | 200×200 | 80×80 | 32 | 3 | 4500 | 80 |
| T2W | 2:07 | N/A | 200×200 | 452×300 | 32 | 3 | 5600 | 170 |

Image processing and data analysis was performed using custom programs written in MATLAB (The MathWorks, Inc). DWI samples acquired at each *b*-value were first averaged together. ADC maps were then computed from the averaged forward-and reverse-polarity DWI volumes by fitting the signal from *b*-values ≤1000 s/mm^2^ with a monoexponential decay model (8, 9, 13, 14).

### Prostate contouring

The prostate contour was automatically defined for each patient on the *T_2_*-weighted volume using a previously validated AI segmentation model (15). A subset of patients also had contours available for target lesions in the prostate that were identified during clinical evaluation and manually defined by an experienced radiologist or radiation oncologist. All contours were converted to binary masks and resampled to the same resolution as the DWI volumes.

### Application of distortion correction methods

For each patient, maps of spatial shift (in units of voxels) were estimated from the forward-and reverse-polarity EPI volumes using RPG, topup, and mRPG. RPG software was provided by request from the developers of the algorithm (4). Topup was obtained as part of the FMRIB Software Library (FSL) (16) version 5.0.2.2, compiled for 64-bit CentOS 6. For both RPG and topup, shift maps were estimated from the *b*=0 s/mm^2^ volumes as described in prior studies, according to the documentation provided with each software package (4–6). The estimated spatial shifts from each method were then applied to the *b*=0 s/mm^2^ images and ADC maps. A separate correction was also performed by applying both the spatial shift maps and JIC to the *b*=0 s/mm^2^ images, to evaluate how the JIC factor specifically influences distortion correction performance. The JIC was not applied to the ADC maps since it is only defined on the domain of MRI signal intensity, not quantitative ADC values.

For mRPG, the forward and reverse DWI volumes were first normalized by their sum across all *b*-values. Spatial shift maps were then computed through registration of the normalized forward-and reverse *b*=0 s/mm^2^ volumes, performed by shifting the normalized volumes along the anterior-posterior axis and identifying the amount of shift for each voxel that minimizes the overall difference between forward and reverse images. A more detailed description of the registration algorithm is provided in the Supplementary Materials.

### Evaluation of anatomical distortion

Extent of anatomical distortion was quantified by the Pearson correlation coefficient between forward and reverse volumes. Scans with minimal distortion will show better alignment between forward and reverse images and a correspondingly higher correlation coefficient than scans with more severe distortion artifacts. Correlation coefficients were computed for the *b*=0 s/mm^2^ and ADC volumes of each patient, before and after distortion correction by each method. Correlation was computed twice per volume, once using the entire field of view (FOV) and once within a prostate region of interest (ROI), defined to include the prostate plus a surrounding 4 mm margin. The size of the margin was adopted from the MIRAGE trial (17), wherein 4 mm margins were applied to account for patient motion and setup uncertainty when creating planning target volumes for CT-guided stereotactic body radiation therapy (SBRT). In the subset of patients with additional DWI repetitions, the correlation was also computed between repeated forward volumes to establish a baseline level of correlation for identical acquisitions that nonetheless could have some variation, e.g., due to patient movement or to physiological prostate motion.

Correlation coefficient distributions across all patients were visualized using violin plots (18, 19). The median and 10^th^ percentile of each distribution was reported with 1000-sample bootstrap 95% confidence intervals. The 10^th^ percentile was chosen to quantify the severity of distortion in patients most impacted by *B_0_*-inhomogeneity artifacts, who have the lowest correlation between forward and reverse volumes. Right-tailed Wilcoxon signed-rank tests (α=0.05) were used to assess whether the Pearson correlation increased significantly after distortion correction by each method, and whether mRPG yielded a larger increase in correlation compared to RPG or topup. The effect size of any changes in correlation before and after distortion correction was assessed using Cohen’s *d*.

Absolute tissue displacement in mm was estimated from the spatial shift maps generated using mRPG. For each patient, the maximum displacement of the posterior prostate border and target lesions was measured. The distribution of maximum tissue displacement across all patients was visualized using a histogram for the prostate border and target lesions, respectively. The median value and range were reported for each distribution. We also reported the number of subjects with maximum tissue displacement exceeding the MIRAGE trial control-arm planning margin of 4 mm, as an indication of the frequency with which distortion artifacts might conceivably impact lesion targeting, were DWI to be used for RT planning. (We note that we use this 4 mm planning margin as a contextual clinical benchmark, rather than as a formally validated threshold for acceptable DWI distortion.)

## Results

### Study participants

In total, 1440 consecutive patients were considered for this study. Of these, 215 were excluded for having implanted metallic devices, yielding 1225 patients for analysis. Target lesion contours were available for 74 patients. Eight patients had their DWI acquisitions repeated within the same scanning session (as participants of a prospective study, enrolled after granting informed consent (7)).

### Evaluation of distortion correction performance

The violin plots in Figure 2 show the distribution of the Pearson correlation between forward and reverse volumes, across all patients. Point estimates and 95% confidence intervals for the median and 10^th^ percentile of each distribution are listed in Table 2. Correlation coefficients measured from repeated forward volumes are shown in Supplemental Figure 1.

**Figure 2:**
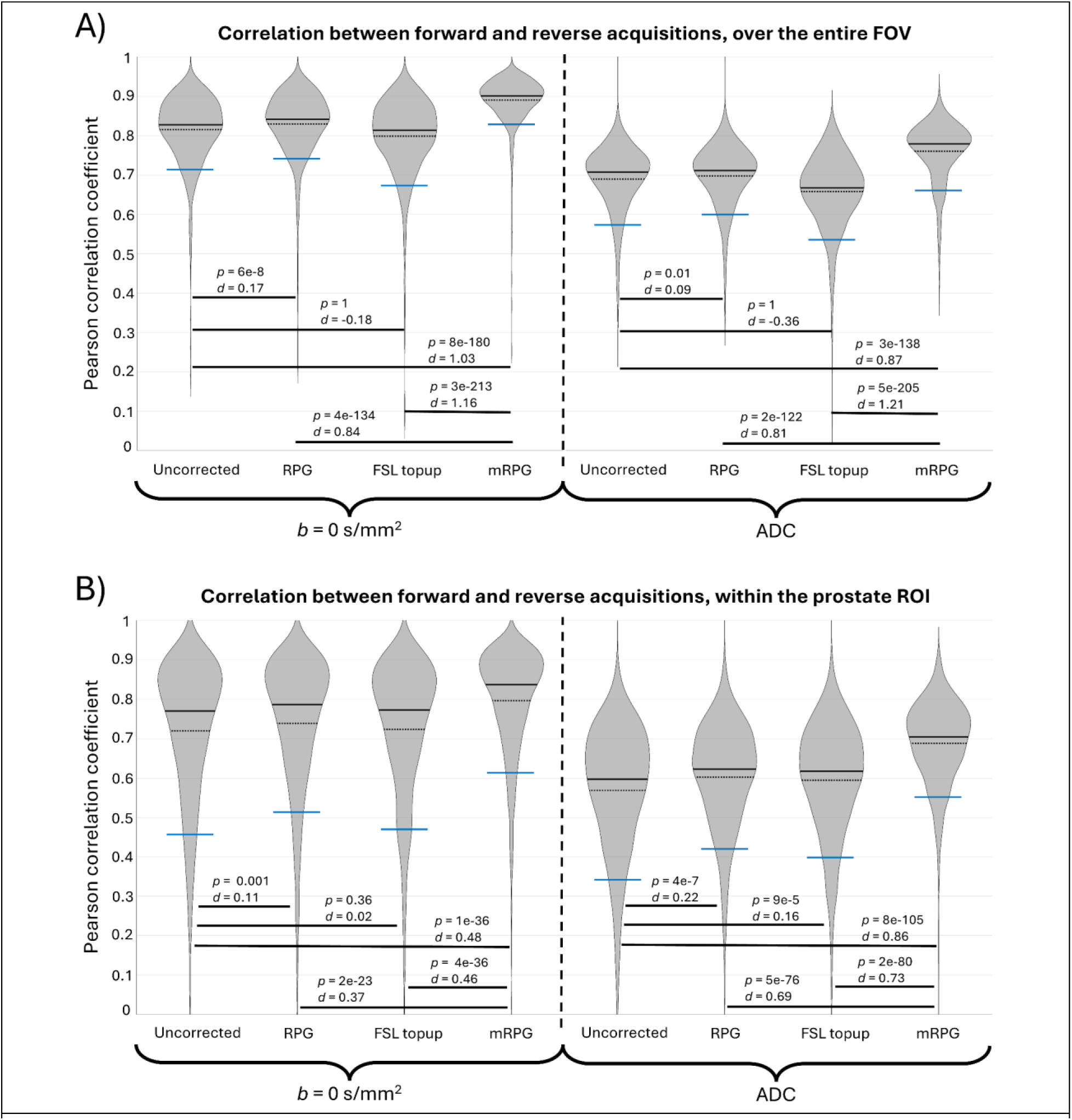
Distribution of correlation coefficients between forward and reverse acquisitions, before and after distortion correction. The 10th percentile, mean, and median of each distribution is indicated by the blue line, dashed black line, and solid black line, respectively. Results of Wilcoxon signed-rank tests and Cohen’s *d* values are indicated on the solid black lines between violin plots. Correlation was measured twice for each *b* = 0 s/mm^2^ and ADC volume, once over the entire field of view (FOV) (A) and once within the prostate region of interest (ROI) (B), defined to include the prostate plus a surrounding 4 mm margin

**Table 2:** Point estimates for the distributions of correlation coefficients shown in Figure 2. The 1000-sample bootstrap 95% confidence intervals are shown in brackets. A) Correlation measured over the entire field of view. B) Correlation measured within the prostate region of interest, defined to include the prostate plus a surrounding 4 mm margin.

| A) | $b = 0 \text{ s/mm}^2$ | | | | ADC | | | |
| --- | --- | --- | --- | --- | --- | --- | --- | --- |
|  | Uncorrected | RPG | FSL topup | mRPG | Uncorrected | RPG | FSL topup | mRPG |
| 10<sup>th</sup> percentile | 0.71<br>[0.70 0.72] | 0.74<br>[0.73 0.75] | 0.68<br>[0.68 0.70] | 0.83<br>[0.82 0.84] | 0.58<br>[0.57 0.59] | 0.60<br>[0.58 0.60] | 0.54<br>[0.53 0.56] | 0.66<br>[0.64 0.67] |
| Median | 0.83<br>[0.82 0.83] | 0.84<br>[0.84 0.84] | 0.81<br>[0.81 0.82] | 0.90<br>[0.90 0.90] | 0.71<br>[0.70 0.71] | 0.71<br>[0.71 0.72] | 0.67<br>[0.66 0.67] | 0.78<br>[0.78 0.78] |

| B) | $b = 0 \text{ s/mm}^2$ | | | | ADC | | | |
| --- | --- | --- | --- | --- | --- | --- | --- | --- |
|  | Uncorrected | RPG | FSL topup | mRPG | Uncorrected | RPG | FSL topup | mRPG |
| 10<sup>th</sup> percentile | 0.46<br>[0.44 0.48] | 0.51<br>[0.48 0.53] | 0.47<br>[0.45 0.49] | 0.61<br>[0.59 0.63] | 0.34<br>[0.32 0.37] | 0.42<br>[0.41 0.44] | 0.40<br>[0.38 0.41] | 0.55<br>[0.54 0.56] |
| Median | 0.77<br>[0.76 0.78] | 0.79<br>[0.78 0.80] | 0.77<br>[0.76 0.78] | 0.84<br>[0.83 0.84] | 0.60<br>[0.59 0.61] | 0.62<br>[0.62 0.63] | 0.62<br>[0.61 0.62] | 0.70<br>[0.70 0.71] |

| **A)** | **$b = 0 \text{ s/mm}^2$** |  |  |  | **ADC** |  |  |  |
| --- | --- | --- | --- | --- | --- | --- | --- | --- |
|  | Uncorrected | RPG | FSL topup | mRPG | Uncorrected | RPG | FSL topup | mRPG |
| 10<sup>th</sup> percentile | 0.71<br>[0.70 0.72] | 0.74<br>[0.73 0.75] | 0.68<br>[0.68 0.70] | 0.83<br>[0.82 0.84] | 0.58<br>[0.57 0.59] | 0.60<br>[0.58 0.60] | 0.54<br>[0.53 0.56] | 0.66<br>[0.64 0.67] |
| Median | 0.83<br>[0.82 0.83] | 0.84<br>[0.84 0.84] | 0.81<br>[0.81 0.82] | 0.90<br>[0.90 0.90] | 0.71<br>[0.70 0.71] | 0.71<br>[0.71 0.72] | 0.67<br>[0.66 0.67] | 0.78<br>[0.78 0.78] |

| **B)** | **$b = 0 \text{ s/mm}^2$** |  |  |  | **ADC** |  |  |  |
| --- | --- | --- | --- | --- | --- | --- | --- | --- |
|  | Uncorrected | RPG | FSL topup | mRPG | Uncorrected | RPG | FSL topup | mRPG |
| 10<sup>th</sup> percentile | 0.46<br>[0.44 0.48] | 0.51<br>[0.48 0.53] | 0.47<br>[0.45 0.49] | 0.61<br>[0.59 0.63] | 0.34<br>[0.32 0.37] | 0.42<br>[0.41 0.44] | 0.40<br>[0.38 0.41] | 0.55<br>[0.54 0.56] |
| Median | 0.77<br>[0.76 0.78] | 0.79<br>[0.78 0.80] | 0.77<br>[0.76 0.78] | 0.84<br>[0.83 0.84] | 0.60<br>[0.59 0.61] | 0.62<br>[0.62 0.63] | 0.62<br>[0.61 0.62] | 0.70<br>[0.70 0.71] |

Application of RPG yielded a significant improvement in correlation between forward and reverse *b*=0 s/mm^2^ volumes, whether measured over the entire FOV (*p* < 0.001, *d =* 0.17) or only within the prostate ROI (*p* = 0.001, *d* = 0.11). Significant improvement was also observed between forward and reverse ADC maps, over the entire FOV (*p* = 0.01, *d* = 0.09) and within the prostate ROI (*p* < 0.001, *d* = 0.22). Topup did not improve the correlation of *b*=0 s/mm^2^ images or ADC maps when computed over the entire FOV (*p* = 1, *d* < 0 for both *b*=0 s/mm^2^ and ADC volumes). In fact, application of topup yielded a lower correlation between forward and reverse volumes than was measured in the images with no distortion correction. Within the prostate ROI, topup produced a statistically significant improvement in the correlation of ADC maps (*p* < 0.001, *d* = 0.16), but not of the *b*=0 s/mm^2^ volumes (*p* = 0.36, *d* = 0.02).

mRPG yielded statistically significant improvements in correlation between forward and reverse *b*=0 s/mm^2^ volumes over the entire FOV (*p* < 0.001, *d* = 1.03) and within the prostate ROI (*p* < 0.001, *d* = 0.48). Improvement in correlation was also significant for the ADC maps over the entire FOV (*p* < 0.001, *d* = 0.87) and within the prostate ROI (*p* < 0.001, *d* = 0.86). The increase in correlation achieved with mRPG was considerably larger than with either RPG or topup for all comparisons (*p* < 0.001, d ≥ 0.37).

**Supplemental Figure 1:**
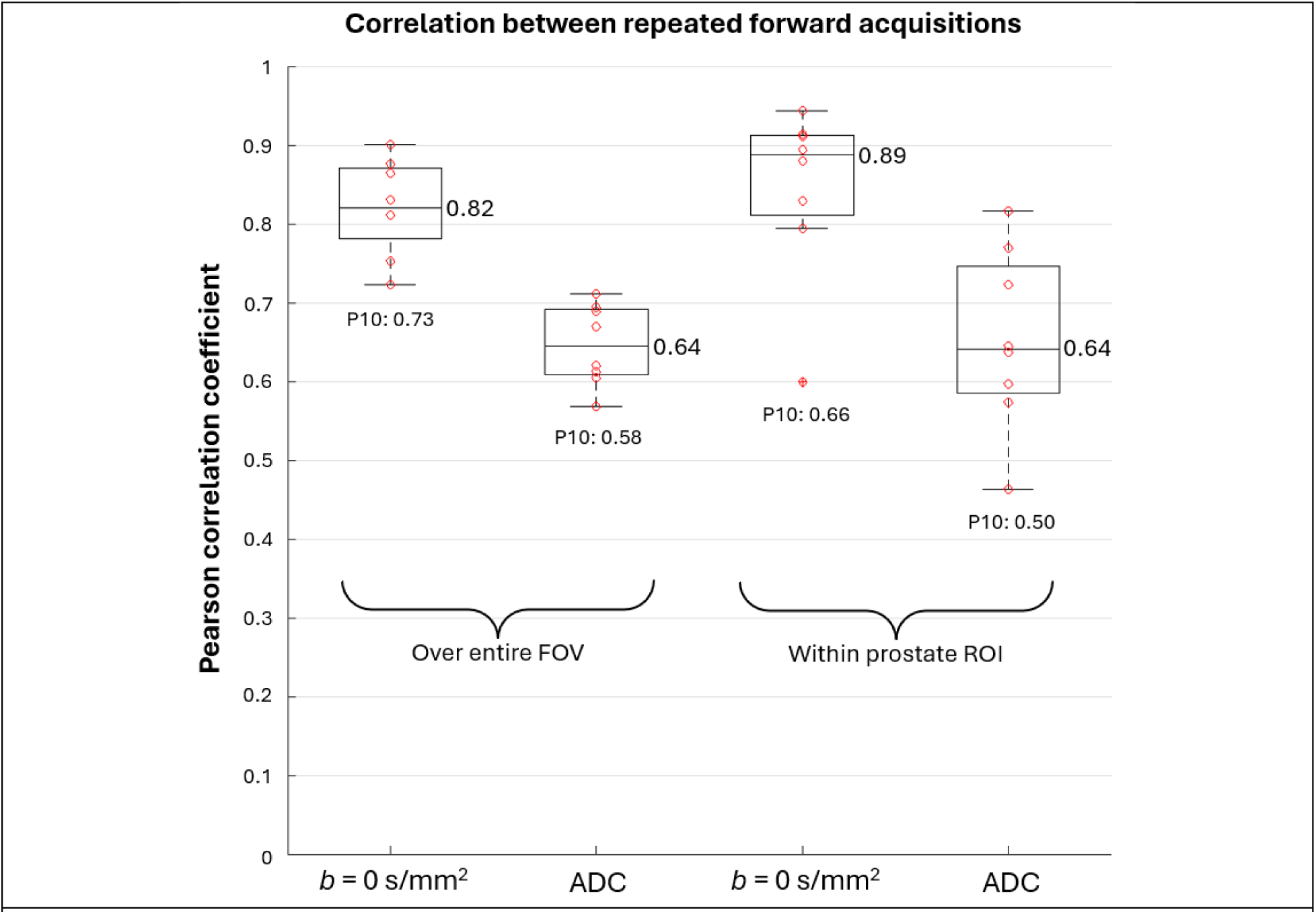
Correlation coefficients measured between repeated forward acquisitions. Measurements from each patient are plotted as red circles over the summary box plots. The median of each distribution is indicated by the central horizontal line, and the 10th percentile (P10) is written below the lowest recorded value. Correlation was measured twice for each *b* = 0 s/mm^2^ and ADC volume, once over the entire field-of-view (FOV) and once within the prostate region of interest (ROI), defined to include the prostate plus a surrounding 4 mm margin. These results demonstrate variation attributable to, for example, patient movement and physiological prostate motion.

### Evaluation of absolute tissue displacement

The histogram in Figure 3A shows the distribution across all patients of the maximum tissue displacement along the posterior border of the prostate, which ranged from 0.5 – 13.8 mm with a median value of 2.5 mm. A maximum displacement ≥ 4 mm (exceeding the MIRAGE trial planning margin) was observed in 30% of patients. The histogram in Figure 3B shows the distribution of maximum tissue displacement within the 74 expert-defined target lesions, which ranged from 0.5 – 7.5 mm with a median value of 1.8 mm.

**Figure 3:**
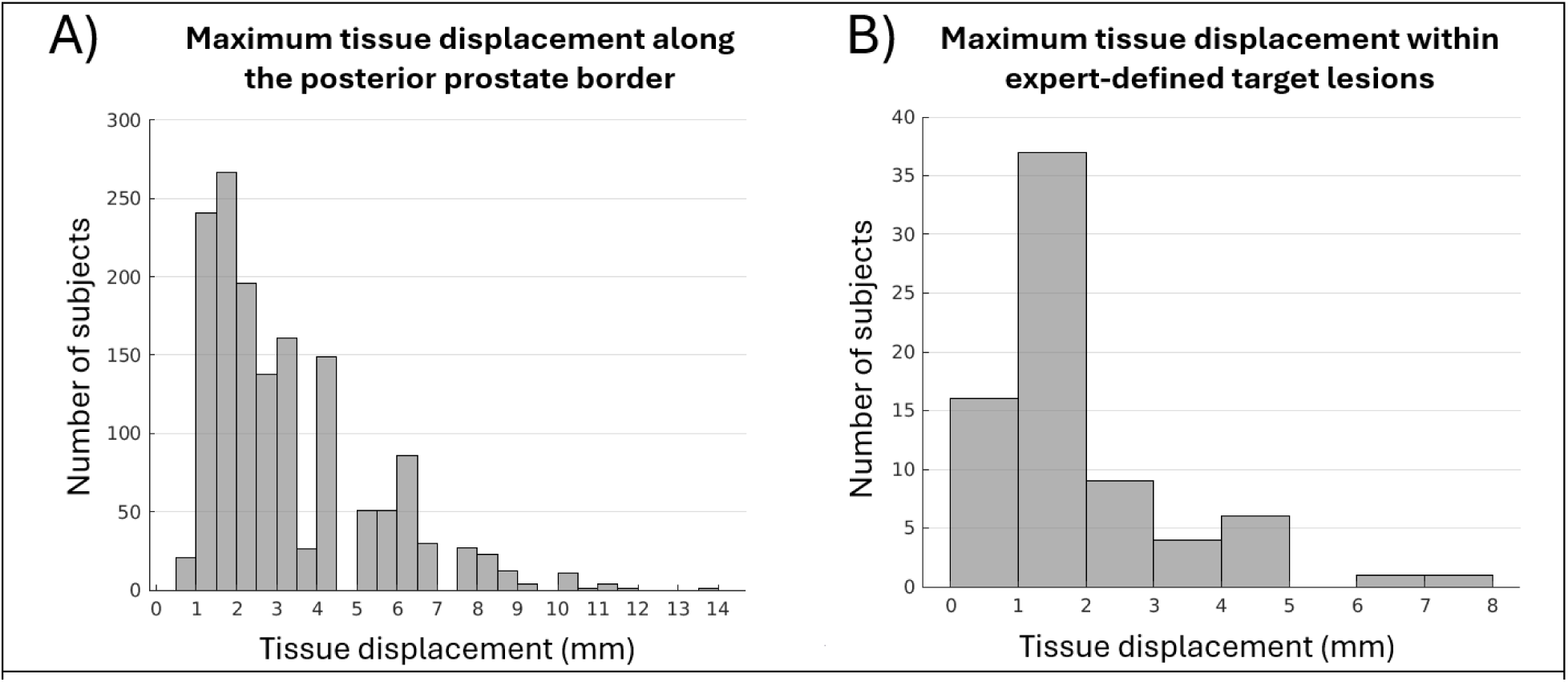
Distribution of maximum tissue displacement in the prostate on DWI. A) Measured along the posterior prostate border of all patients. B) Measured within 74 expert-defined target lesions

### The impact of the Jacobian Intensity Correction (JIC) on distortion correction performance

The violin plots in Figure 4 show the distribution across all patients of the Pearson correlation between forward and reverse volumes, with and without JIC applied for RPG and topup. With JIC applied, both RPG and topup yielded a considerable increase in correlation between forward and reverse *b*=0 s/mm^2^ acquisitions, with median correlation ≥ 0.95. This increase cannot be attributed to an accurate estimation of absolute tissue displacement, however, since no parallel increase in correlation was observed when the estimated spatial shifts were applied without the JIC. This disparity is most apparent with topup, which showed a *decrease* in correlation after application of the spatial shift maps alone.

**Figure 4:**
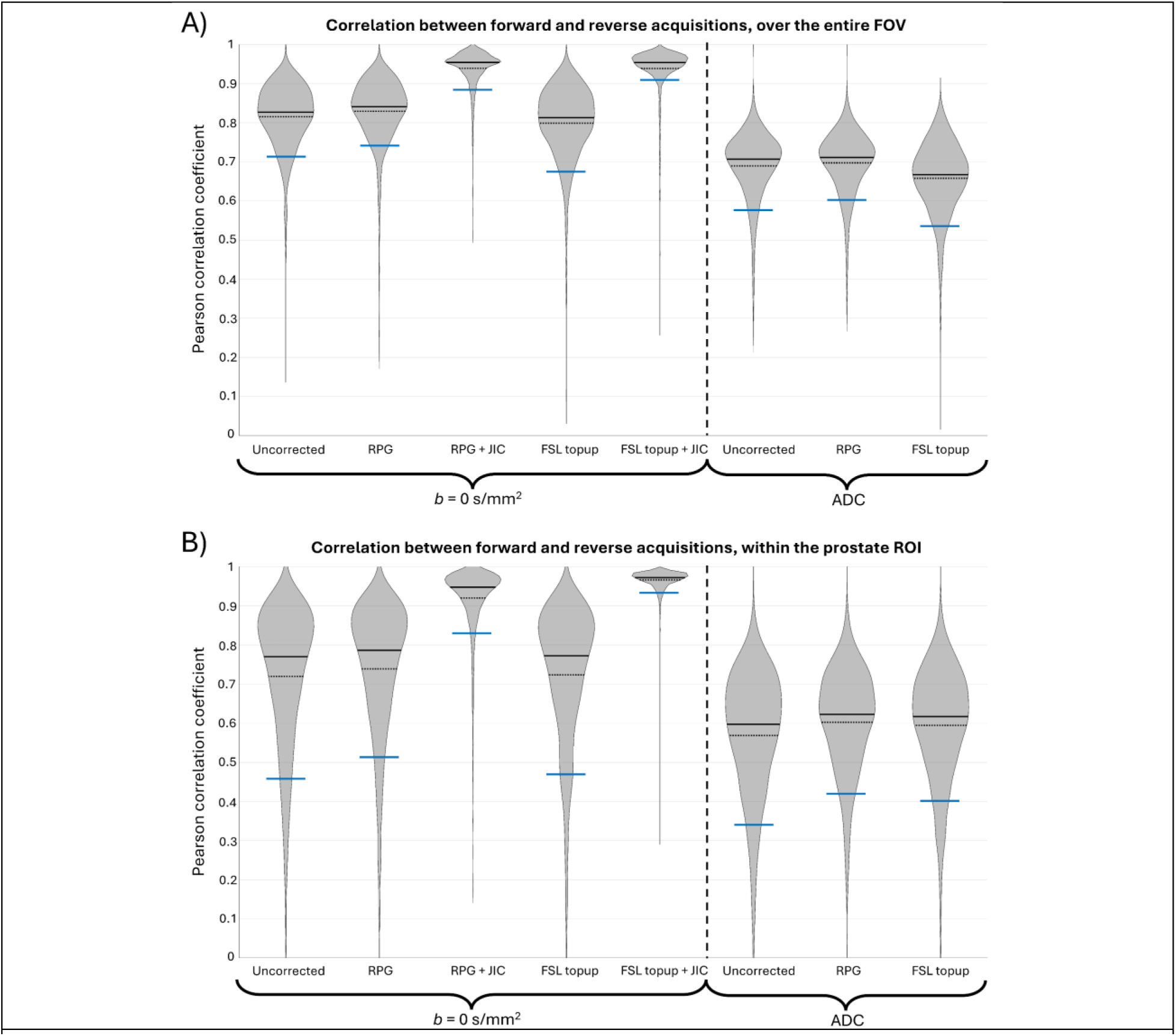
Correlation between forward and reverse acquisitions, before and after distortion correction that includes application of the Jacobian Intensity Correction (JIC). The 10th percentile, mean, and median of each distribution is indicated by the blue line, dashed black line, and solid black line, respectively. Correlation was measured twice for each *b* = 0 s/mm^2^ and ADC volume, once over the entire field of view (FOV) (A) and once within the prostate region of interest (ROI) (B), defined to include the prostate plus a surrounding 4 mm margin. RPG and topup only yielded substantial increases in the correlation of *b*=0 s/mm^2^ images when the JIC was applied. Improvement without the JIC, if any, was considerably smaller. Such a disparity suggests an underestimation of absolute tissue displacement with RPG and topup that is concealed by the intensity changes applied by the JIC. This underestimation is also evident in the corrected ADC maps, which similarly show only a marginal increase in correlation compared to the much larger improvement observed on *b* = 0 s/mm^2^ images with the JIC.

In other words, application of the JIC has the potential to conceal poor estimation of tissue displacement by artificially adjusting signal intensity. An example of such misleading concealment of poor distortion correction is illustrated in Figure 5. While the uncorrected diffusion images from this patient showed minimal distortion, both RPG and topup incorrectly estimated displacement of the anterior and posterior regions of the prostate. Overcorrection is readily apparent when the estimated spatial shifts are applied without the JIC, particularly for RPG. Application of the JIC obscured the overcorrection, however, adjusting image intensity to yield an apparent alignment between the EPI images and the reference spin echo image. For RPG, the JIC also resulted in false anatomical structure or “ghost tissue” between the posterior prostate and rectum. For topup, the JIC reduced the signal intensity in a portion of the posterior prostate in the forward image to match the signal intensity of the rectum in the reverse image. mRPG, by contrast, yielded appropriate spatial correction for this patient.

**Figure 5:**
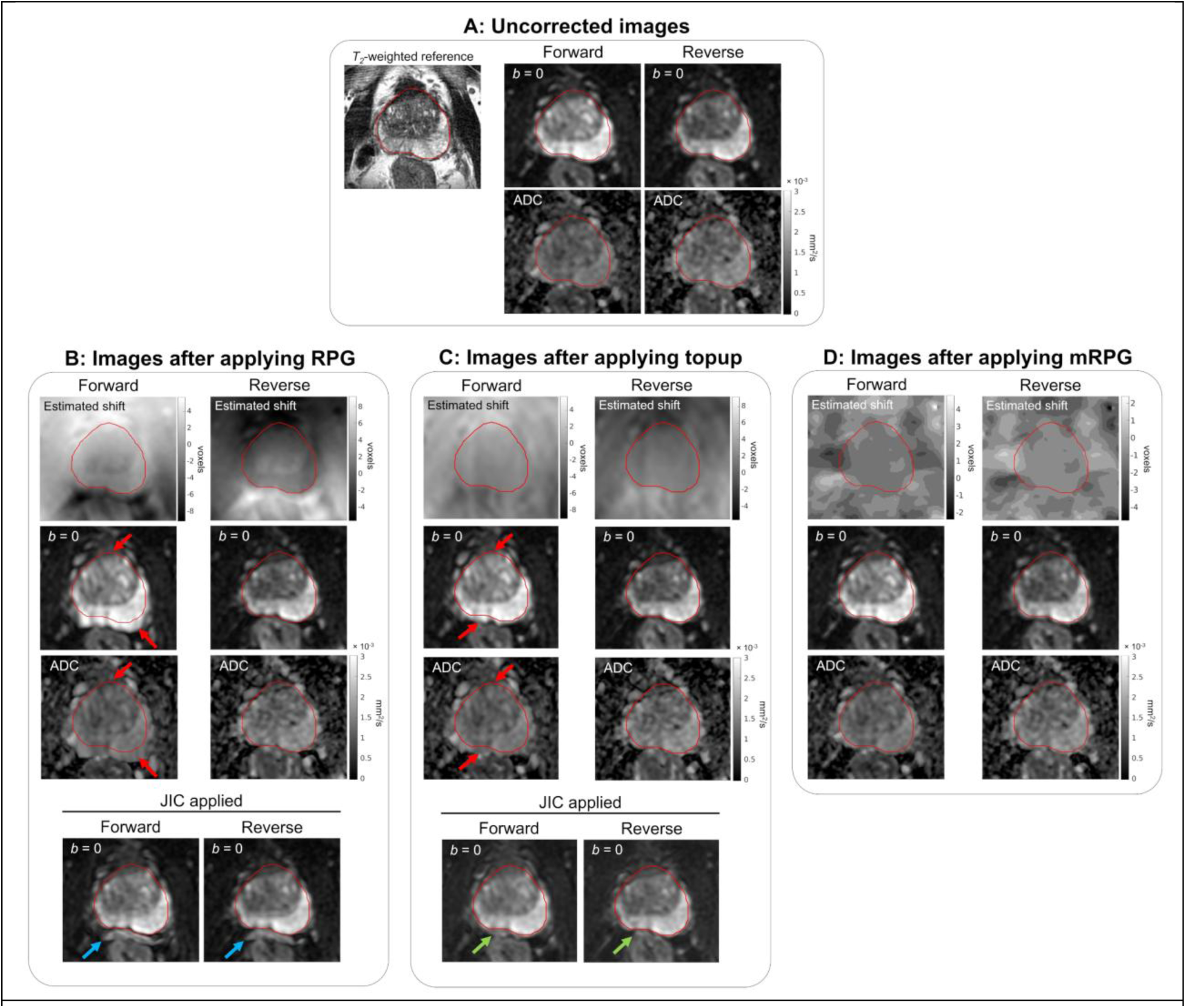
Example of the Jacobian intensity correction (JIC) concealing poor estimation of tissue displacement. A) An undistorted spin echo image is shown with the prostate outlined in red. Uncorrected diffusion images show minimal B0-inhomogeneity distortion. RPG (B) and topup (C) incorrectly overestimated shifts in the anterior and posterior prostate (red arrows). JIC concealed the erroneous spatial shift while also creating false anatomical structure or “ghost tissue” (B, blue arrows), and reducing the signal intensity of a portion of the prostate to match that of the rectum (C, green arrows). mRPG, by contrast, yielded appropriate spatial correction (D).

## Discussion

The mRPG approach for correction of *B_0_*-inhomogeneity distortion outperformed conventional methods. mRPG yielded markedly higher correlation between forward-and reverse-phase-encoding polarity images than either RPG or FSL topup, indicating a greater reduction in anatomical distortion. Effective correction of distortion artifacts is particularly important in the context of focal RT boost to MRI-visible tumors, which requires high accuracy to ensure delivery of ablative RT doses while avoiding excess dose to neighboring organs of interest (20). We found that prostate tissue displacement due to *B_0_*-inhomogeneity is frequently greater than 4 mm (the SBRT planning margin used for the MIRAGE trial), and can be more than a centimeter in severe cases. Distortion correction with a technique like mRPG is necessary to optimize targeting accuracy when using DWI or ADC.

While the correlation between forward and reverse acquisitions after correction with mRPG was not perfect, it was comparable to the correlation between repeated acquisitions of the same phase-encoding polarity. This result suggests that differences between forward and reverse images that remain after correction with mRPG are largely due to factors other than *B_0_*-inhomogeneity distortion. Much of the pelvic FOV is comprised of fatty tissue with minimal signal intensity due to the fat-signal suppression used during image acquisition, resulting in many areas that are essentially random noise patterns (even without diffusion weighting). These regions of random noise lower the maximum achievable correlation between images, even if patient anatomy is perfectly identical between acquisitions. Other anatomical factors like gross patient motion and soft tissue deformation from bladder filling and rectal gas flow can also limit the correlation between forward and reverse images. With these considerations in mind, mRPG offers the best correction for *B_0_*-inhomogeneity distortion out of the three methods examined in this study.

RPG and topup yielded more modest changes to image correlation. Notably, however, the application of topup actually *reduced* the overall correlation between forward and reverse images. Visual inspection of images corrected with topup suggested that this reduction was in large part a result of erratic tissue deformations applied in the periphery of the field of view. Erroneous deformations applied to peripheral tissues are less concerning than distortion within the prostate, but might still warp the anatomy of structures like lymph nodes and bones that are routinely assessed for metastatic disease (21, 22). Within the prostate ROI, topup and RPG both reduced distortion to similarly modest degree, though neither performed as well as mRPG.

While the tissue deformation fields estimated with RPG and topup produce only slight changes in the correlation between forward and reverse images, application of the JIC results in striking improvement. Median correlation between forward and reverse *b*=0 s/mm^2^ images after JIC was considerably higher than between repeated forward acquisitions, which are generally subject to the same *B_0_*-inhomogeneity distortions. It is apparent from the results observed without the JIC, however, that the improvement afforded by the JIC is divorced from the accuracy of underlying tissue deformation estimates and can even conceal glaring inaccuracies. The disparity observed in distortion correction performance with and without the JIC suggests an overfitting problem during the minimization of Eq. 3, where the solution does not depend on finding the absolute tissue displacement that yields the best alignment between images, but rather which voxel-wise changes to signal intensity yield the highest overall similarity. mRPG circumvents this overfitting problem by first eliminating the signal-intensity scaling factor and only then estimating tissue deformation.

For this study, we acquired approximately 3 minutes of additional reverse-polarity images for mRPG to achieve improved accuracy. Our acquisition was longer than strictly necessary because our acquisition protocol was aligned with a prospective trial (NCT06579417) that is conservatively optimized for higher signal-to-noise ratio (SNR) for the DWI data (11). In principle, only two unique *b*-values are required in the reverse phase-encoding direction to perform the image normalization required for mRPG, and we have separately employed such an abbreviated approach for breast imaging (23). However, repeating the full acquisition in both directions does provide an SNR advantage since the full set of images in both directions can be averaged together after distortion correction. We do recommend that the same *b*-values are used for normalization in both phase-encoding directions so that tissue contrast in the normalized images is consistent between the two. If in this study, for example, only *b* = 0 and 800 s/mm^2^ were acquired in the reverse direction, we would have similarly computed the normalized image volume in the forward direction using only *b* = 0 and 800 s/mm^2^, rather than the whole series of 5 *b*-values.

A limitation of this study is that the efficacy of distortion correction was only measured using the correlation between opposite-polarity images. While correlation provides a quantitative measure of the alignment between images, it is not an assessment of diagnostic image quality *per se*. A phantom study could measure distortion more precisely but would not reflect the range of physiological possibilities encountered in clinical practice. Meanwhile, the clinical value of mRPG in a diagnostic radiology setting is currently being assessed by expert radiologists as a secondary endpoint of a multi-center, international clinical trial (NCT06579417) (11). The use of mRPG-corrected images for pre-biopsy imaging and tumor-focused RT are also being evaluated in separate randomized trials (NCT05882253, NCT06990542) (12).

## Conclusion

mRPG is a feasible method for correcting *B_0_*-inhomogeneity distortion artifacts in prostate DWI data. It avoids significant drawbacks of alternative methods to achieve better distortion correction performance, which might be useful in diagnostic imaging, MRI-guided biopsy, and/or targeting for focal RT boost.

## Data Availability

All data produced in the present study are available upon reasonable request to the corresponding author

## Supplementary Materials

### *B_0_*-inhomogeneity distortion correction algorithm

- Inputs: Two sets of multi-b-value DWI images acquired using EPI readouts of opposite phase-encoding polarity, arbitrarily referred to as the “forward” and “reverse” sets of images.
- Output: A voxel-wise map of displacement (in units of voxels) that can be applied to the forward set of DWI images to correct *B0*-inhomogeneity distortions. The negative of the displacement map can be applied to correct the reverse set of images.

### DWI volume preprocessing

1. Averaged DWI image volumes are first computed for the forward and reverse image sets by taking the arithmetic mean of all samples (directions/repetitions) acquired at each b-value
2. The averaged forward and reverse DWI volumes are then normalized by the sum across all b-values
3. The averaged, normalized forward and reverse *b*=0 s/mm^2^ volumes are returned as the inputs for the subsequent registration algorithm

### Registering normalized forward and reverse *b*=0 s/mm^2^ volumes

1. The normalized volumes are smoothed via Gaussian blur. In this study, **σ** was set to 10/(voxel size along each dimension). Most DWI images had voxel dimensions of [1.25, 1.25, 3] mm in the row, column, and slice direction, respectively, yielding **σ** of [8, 8, 3.33]
2. Initialize the displacement map (*Di*) to zero for all voxels in the field of view
3. Iterate through a range of possible displacement magnitudes, *d*, from −40 to +40 voxels along the phase-encoding direction in increments of 0.8 voxels (for a total of 101 different values of *d*):

I. Shift the forward volume by +d along the phase encoding direction
II. Shift the reverse volume by -d along the phase encoding direction
III. Compute the value of the cost function for all voxels:

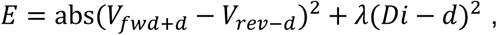

where *E* is the volume of cost function values, *V_fwd+d_* and *V_rev-d_* are the forward and reverse volumes, respectively, shifted along the phase encoding direction, *λ* is the regularization parameter, *Di* is the current estimate for the displacement map, and *d* is the current displacement magnitude. The first term of the cost function is minimized when the shifted forward and reverse volumes are aligned, while the regularization term constrains the magnitude of estimated voxel displacements. In this study, *λ* was set to 1e-4 for the first iteration of the algorithm and 1e-3 for all subsequent iterations.
IV. The cost volume *E* is smoothed using robust spline smoothing (1, 2) with the smoothing parameter set to 1.
4. For each voxel, the value of *Di* is updated to the magnitude *d* which produced the smallest value of *E*
5. Repeat steps 3 and 4 until convergence. In this study, 3 repetitions were used for all registrations.

1. Garcia D: Robust smoothing of gridded data in one and higher dimensions with missing values. *Computational Statistics & Data Analysis* 2010; 54:1167–1178.

2. Garcia D: A fast all-in-one method for automated post-processing of PIV data. *Exp Fluids* 2011; 50:1247–1259.

